# Burden of Human Metapneumovirus-associated pneumonia among children attending a Tertiary Care Hospital, Kathmandu

**DOI:** 10.1101/2022.09.04.22279574

**Authors:** Jyoti Lamichhane, Milan Upreti, Krishus Nepal, Bishnu Prasad Upadhyay, Urusha Maharjan, Ram Krishna Shrestha, Ram Hari Chapagain, Megha Raj Banjara, Upendra Thapa Shrestha

## Abstract

**Background:** Acute Respiratory Tract Infection (ARI) is one of the most common causes of mortality and morbidity worldwide. Every year millions of children suffer from viral Respiratory Tract Infections (RTIs) ranging from mild to severe illnesses. Human Metapneumovirus (HMPV) is among the most frequent viruses responsible for RTIs. However, HMPV infections and their severity among children have not been explored yet in Nepal. Therefore, the study aimed to determine the incidence of HMPV among children attending Kanti Children’s Hospital and assess the clinical characteristics of the infections as well as find out the co-infections.

**Methods:** A hospital-based cross-sectional study was carried out from September 2018 to April 2019. A total of 105 throat swabs were collected from children clinically suspected of ARIs. The collected samples were tested for the presence of HMPV RNA by Multiplex Real-time PCR (RT-PCR) assay.

**Results:** The incidence of HMPV in children in Kanti Children’s Hospital was found to be 13.3%. The HMPV was more prevalent in the age group less than three years (21.8%) which was found to be statistically significant (p - 0.038). Cough and fever were the most common clinical features present in all children infected with HMPV followed by rhinorrhea, sore throat, and wheezing. HMPV positive children were diagnosed with pneumonia (42.9%), bronchiolitis (28.5%), upper respiratory tract infections (14.3%) and asthma (14.3%). The incidence of HMPV was high in late winter (14.3%) followed by early spring (13.5%).

**Conclusion:** This study provides the baseline information on HMPV and associated co-infection with other respiratory viruses for differential diagnosis and rational use of antibiotics in clinical management.

## INTRODUCTION

Acute Respiratory tract infections (ARTIs) pose a major public health problem worldwide with significant morbidity and mortality (Kahn 2006, Panda et al 2014). Annually, 4.3 million people suffer from acute respiratory tract infections globally (Mabey et al 2004). About 13 million children < 5 years of age, die every year, 95% of them from developing countries and one-third of the total deaths are due to ARIs (Prajapati et al 2011). Pneumonia and bronchiolitis are the leading contributors to the global burden of ARIs in young children. These infections are responsible for the greater part of the mortality rate, of which the vast majority occur in developing countries. Pneumonia alone kills a greater number of children than any other illness: AIDS, malaria, and measles combined (Mathisen et al 2010).

Viral agents are among the most common pathogens responsible for respiratory tract infections in young children (van Woensel et al 2003; Haas et al 2013). However, the etiological agents for a large number of respiratory tract infections (RTIs) remain unknown (Bastien et al 2003). Traditionally, respiratory syncytial virus (RSV), parainfluenza virus (PIV), influenza virus, and adenovirus have been considered the leading causes of acute viral respiratory tract infections (Debiaggi et al 2012). Since the advent of more sensitive diagnostic tools, like PCR, the proportion of known viral etiologies has increased. In the last ten years due to the advancement in molecular technologies, newly discovered viruses have been identified from patients of RTIs, like human Metapneumovirus, Coronaviruses NL63, and HKU, Human Bocavirus, new Enterovirus, Parechovirus, and Rhinovirus strains, Polyomaviruses WU and KI and the pandemic H1N1 influenza A virus (Debiaggi et al 2012).

Infections with HMPV can be both symptomatic and asymptomatic (Arnott et al 2013). Clinical manifestations of symptomatic HMPV are indistinguishable from those of RSV and range from mild upper respiratory tract infection to severe diseases requiring hospitalization like severe cough, bronchiolitis, and pneumonia, often accompanied by high fever, myalgia, and vomiting (Schildgen et al 2011). The incubation period is estimated to be 3 to 6 days, and the median duration of illness can vary depending upon severity but is similar to other respiratory infections caused by other viruses (CDC 2016). Re-infection with HMPV is common (Arnott et al 2013). To date there is no vaccine is available (CDC 2016).

HMPV can infect people of all age groups, with a high prevalence in pediatric patients. The first HMPV infection appears to take place at 6 months of life, after which infections may occur repeatedly and frequently. The elderly represent the second group of patients that are severely affected by HMPV (Miroballi et al 2010). About 38% of elderly patients with HMPV infections used medical care in contrast to 9% of young HMPV-infected patients (Haas et al 2013). HMPV is also a significant cause of acute respiratory diseases in adults with comorbid diseases, such as COPD, asthma, cancer, and immune-compromised status, including HIV or post-transplantation (Falsey et al 2003). HMPV is found to be associated with a severe infection in patients with pulmonary disease and chronic obstructive pulmonary disease (COPD) (Haas et al 2013).

HMPV infections can occur throughout the year, but seasonality has been described in several studies (Nidaira et al 2012, Horton et al 2017). The seasonal distribution of HMPV was found to be largely similar to that of RSV, with the peak of virus detection in winter (van den Hoogen 2003). However, it varies from year to year and place to place (Debiaggi et al 2012).

Since the advancement of molecular techniques, the detection of co-infections by multiple respiratory viruses from the same respiratory specimen has been made possible. In children hospitalized due to severe bronchiolitis, coinfection may reach 70% according to some reports, although most studies have shown that prevalence rates range from 15 to 39% (Semple et al 2005; Calvo et al 2008). Because Human metapneumovirus is relatively new and not well described, the chances of underdiagnosis in regular medical practice are very high. Center for Disease Control and Prevention (CDC) recommends considering metapneumovirus testing along with Influenza virus (Flu), RSV, and other common respiratory viruses, especially in patients with severe respiratory illness. Despite that, there are only limited reports regarding HMPV infections. Studies had shown that HMPV is one of the prevalent causes of respiratory tract infection in south Asian countries including Nepal (Mathisen 2010; Lenahan et al 2017). Dr. Mathisen (2010) found that about 4.2% of pneumonia in children in Bhaktapur, Nepal was caused by HMPV. Women of Nepal with HMPV during pregnancy had an increased risk of giving birth to infants who were small for gestational age (Lenahan et al 2017).

Actual data regarding the prevalence of HMPV alone in our country is not known however some studies had described the co-infection of HMPV with other viruses (Upadhyay et al 2018) and bacteria (Murray et al 2018). In addition to the limited data regarding the epidemiology of HMPV in children in subtropical regions such as Nepal; the seasonal pattern remains unknown. Many studies also lack the correlation of clinical syndromes with HMPV infections. On the other hand, using multiplex real-time PCR for diagnosis is rarely used in our contest, the study aimed to focus on the diagnosis of potential viral etiologies and coinfections among children using multiplex PCR visiting a tertiary care hospital in Kathmandu, Nepal.

## METHODS

### Study approval and consent

The study was approved by the review committee, Kanti Children’s Hospital, Maharajgunj, Kathmandu, Nepal (Reference no.: 2018019). After giving brief information about this research, written informed consent and assent were obtained from respective parents/guardians and children prior to sample collection.

### Inclusion and exclusion criteria

#### Study Population

Children below 15 years suspected of influenza-like illness with respiratory tract infection were enrolled in the study. A total of 105 patients from Kanti Children’s Hospital, Maharajgunj, Kathmandu were included in the study. Among them, 90 were from the outpatients’ department and 15 were admitted to the ward.

#### Data collection

The Clinical history of HMPV-infected children was obtained from pediatricians’ notes. Clinical data included demographic data (sex, age, and underlying disease of the patient), clinical symptoms (cough, rhinitis, body temperature, dyspnea, wheezing, feeding difficulties, retractions, Headache), and clinical diagnosis (URTI, Pneumonia, Bronchiolitis, Asthma).

#### Sample collection

A total of 105 non-duplicate throat swabs samples were collected using a standard microbiological technique for three fixed days, every week from January 2019 to March 2019 (Health Protection Agency, 2012). The samples were collected with a clean, dry and sterile Dracon swab stick, which was then immediately kept in a tube containing Viral Transportation Media (VTM) and transported to Central Diagnostic Laboratory and Research Center, Kamalpokhari, Kathmandu. Samples were stored at -20 °C until further processing.

### Extraction and purification of viral nucleic acids

RNA was isolated and purified from a 200 µl cell-free sample by using the PureLink™ Viral RNA/DNA Mini kit, the entire procedure was carried out according to kit instructions. The viral particles in the cell-free samples are lysed using Proteinase K and Lysis Buffer (L22) containing 5.6 μg Carrier RNA at 56°C. The Lysis Buffer (L22) was specifically formulated to allow the lysis of different types of viral particles. The presence of an excess amount of Carrier RNA (yeast tRNA) in the lysate preparation and purification process increased the binding of viral nucleic acids to the silica matrix and reduced any viral nucleic acid degradation from nucleases present in the sample. Ethanol was added to the lysate to a final concentration of 37% and the sample was loaded onto a silica spin column. The viral RNA/DNA molecules bound to the silica-based media and impurities such as proteins and nucleases are removed by thorough washing with Wash Buffer. The RNA/DNA was then eluted in sterile, RNase-free water.

### Polymerase Chain Reaction

All the throat swab samples were tested for HMPV by real-time PCR. The viral RNA is transcribed into cDNA using a specific primer-mediated reverse transcription step followed immediately in the same tube by a polymerase chain reaction. The presence of specific viral sequences in the reaction is detected by an increase in fluorescence observed from the relevant dual-labeled probe and is reported as a cycle threshold value (Ct) by the Real-Time thermocycler. The preparation of PCR was done with a master mix (Fast Tract Diagnostica, Luxemburg, Finland). Reagents for the reaction: BoMpPf1 PP, the positive control (PC), and 2x RT-PCR buffer (light blue cap) were thawed completely and the reaction mix was prepared as shown in Table 1. Then 96 well plate compatible with the Cfx 96, Bio-Rad, USA was taken; 15μl of the reaction mix (BoMpPf1 PP) was pipetted in the wells. 10μl of the extracted samples, the extracted negative control, and the positive control were added in wells and labeled correctly. Each run included a negative and positive control. The reaction mix with samples/PC/NC was mixed well by pipetting up and down. The plate was closed with the ABI optical adhesive film and briefly centrifuged afterward. Then, the plate was put in real-time thermal cycler Cfx 96, Bio-Rad, USA.

**Table 1:**
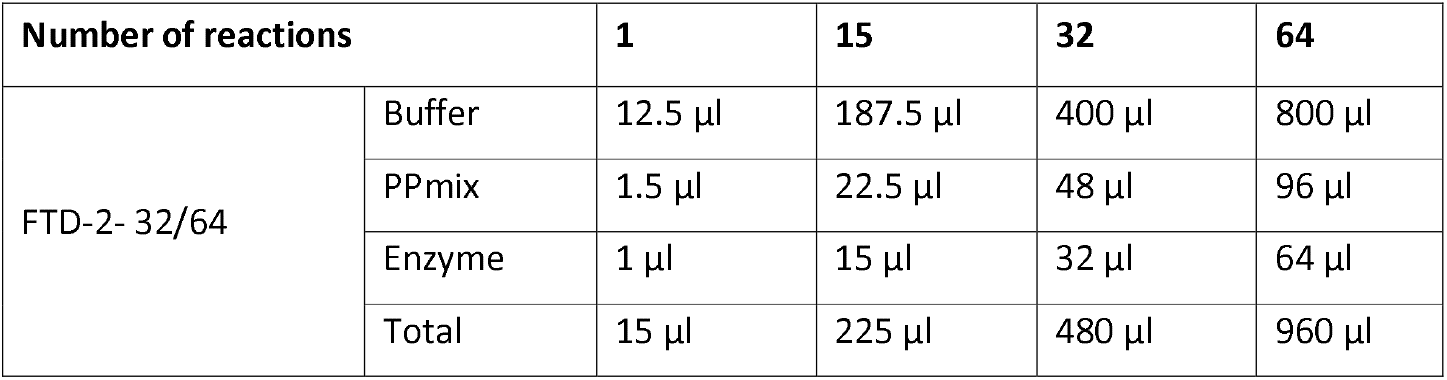
The amounts of reagents needed for 1, 15, 32, and 64 wells.

| Number of reactions |  | 1 | 15 | 32 | 64 |
| --- | --- | --- | --- | --- | --- |
| FTD-2- 32/64 | Buffer | 12.5 µl | 187.5 µl | 400 µl | 800 µl |
|  | PPmix | 1.5 µl | 22.5 µl | 48 µl | 96 µl |
|  | Enzyme | 1 µl | 15 µl | 32 µl | 64 µl |
|  | Total | 15 µl | 225 µl | 480 µl | 960 µl |

Particular attention was paid to the settings for the detectors (Table 2). RT-PCR Amplification was performed with the following cycling parameters: 42°C for 15 minutes hold; 94°C for 3 minutes hold; 40 cycles of 94°C for 8 seconds and 60°C for 34 seconds.

**Table 2:**
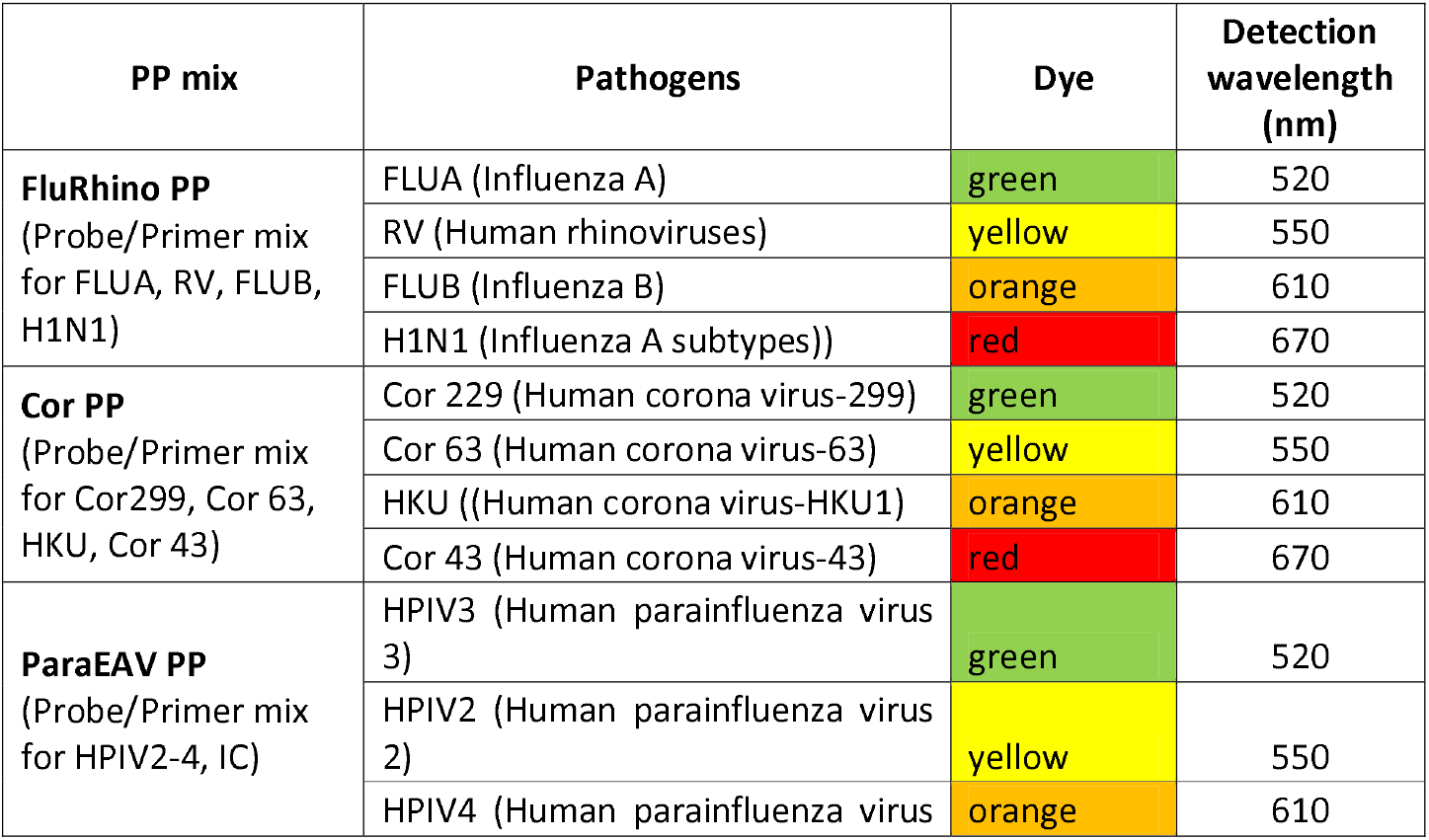
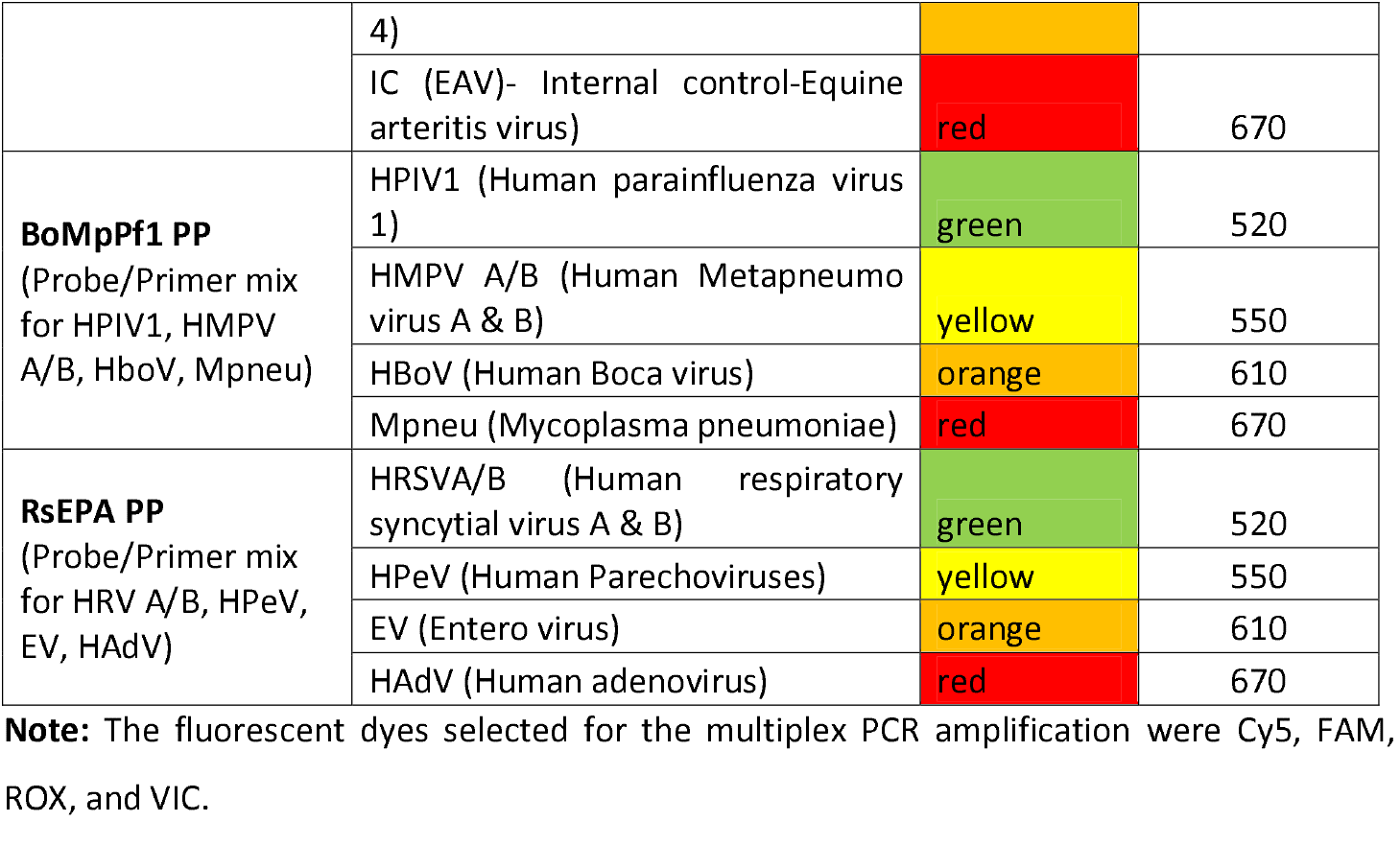
Settings of the detectors for detection of multiple pathogens.

### Quality control

A threshold was set according to manufacturer instructions. All negative controls were below the threshold and positive control showed the positive (i.e exponential) amplification curve. Internal control also showed the positive (i.e exponential) amplification tract as well. Therefore, the process is valid. After all, the controls met the specified ranges, and all samples were checked for positive traces. Ct results for all color channels were displayed on the “View Well Table” window.

### Data Analysis

All the data were entered and analyzed by using Statistical Package for Social Science (SPSS) version 24 software package. The result having a p-value < 0.05 was considered significant.

## RESULTS

### Incidence of HMPV

Out of 105 specimens collected from children with ARIs, 14 (13.3%) were positive for HMPV by multiplex-PCR. The incidence of HMPV among children was observed to be 13.3% (95% CI: 7.0-20). The positive rate of HMPV in females was 13% and the male was 13.3%. HMPV-infected children were aged from newborn to 14 years (median age: 4 years). Most infected individuals were from the outpatient department (Table 3).

**Table 3:** Incidence of HMPV in the study population.

| Distribution of HMPV |  | Total no. | No. of HMPV positive cases (%) | No. of HMPV negative cases (%) | p-value |
| --- | --- | --- | --- | --- | --- |
| Gender | Female | 46 | 6 (13) | 40 (87) | >0.05 |
|  | Male | 59 | 8 (13.5) | 51 (86.5) |  |
| Age Group (yrs) | <3 | 32 | 7 (21.8) | 25 (78.2) | 0.038 |
|  | 3-5 | 45 | 6 (13.3) | 39 (86.7) |  |
|  | 6-15 | 28 | 1 (3.5) | 27 (96.5) |  |
| Patient Type | Out-patient | 90 | 12 (11.4) | 78 (88.6) | >0.05 |
|  | In-patient | 15 | 2 (1.9) | 13 (98.1) |  |

### Clinical characteristics of a patient infected with HMPV

The most common clinical findings of HMPV infection were cough (14; 100%) and fever (14; 100%) followed by Rhinorrhea (10, 71.4%), sore throat (9; 64.3%), and wheezing (7; 50%). Three of these patients (3; 21.4%) had complaints of loss of appetite and one (1; 7.1%) had a headache. Out of 14 HMPV-positive cases, 42.9% (6) of the patients were diagnosed with pneumonia. Similarly, 28.5% (4) with bronchiolitis, 14.3% (2) diagnosed with URT, and 14.3% (2) with asthma (Table 4).

**Table 4:**
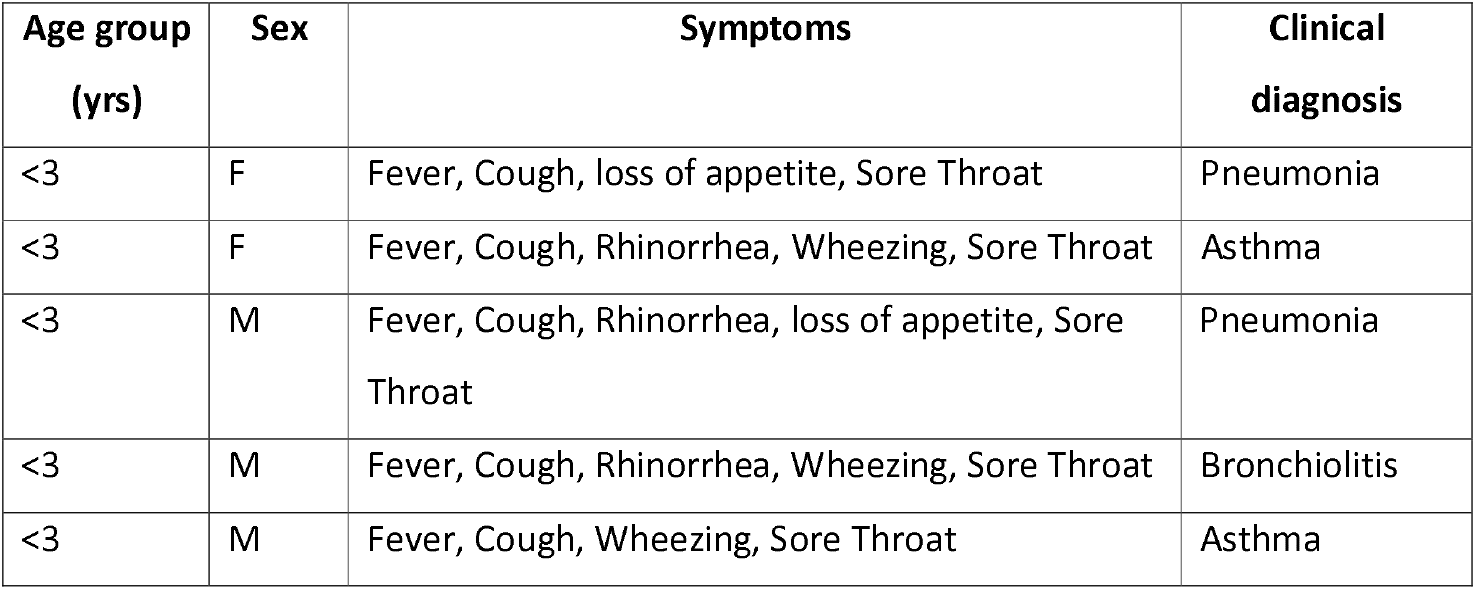
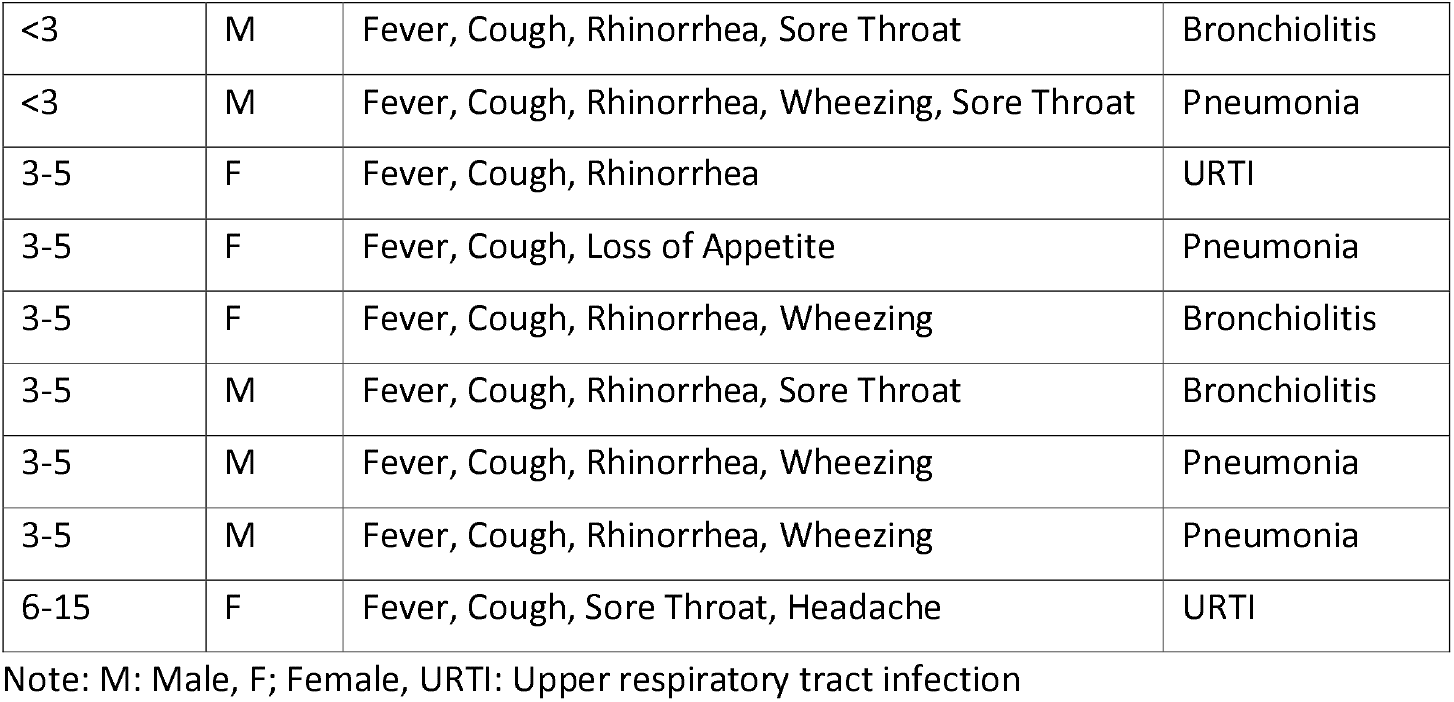
Clinical characteristics of HMPV-positive patients.

| Age group (yrs) | Sex | Symptoms | Clinical diagnosis |
| --- | --- | --- | --- |
| <3 | F | Fever, Cough, loss of appetite, Sore Throat | Pneumonia |
| <3 | F | Fever, Cough, Rhinorrhea, Wheezing, Sore Throat | Asthma |
| <3 | M | Fever, Cough, Rhinorrhea, loss of appetite, Sore Throat | Pneumonia |
| <3 | M | Fever, Cough, Rhinorrhea, Wheezing, Sore Throat | Bronchiolitis |
| <3 | M | Fever, Cough, Wheezing, Sore Throat | Asthma |
| <3 | M | Fever, Cough, Rhinorrhea, Sore Throat | Bronchiolitis |
| <3 | M | Fever, Cough, Rhinorrhea, Wheezing, Sore Throat | Pneumonia |
| 3-5 | F | Fever, Cough, Rhinorrhea | URTI |
| 3-5 | F | Fever, Cough, Loss of Appetite | Pneumonia |
| 3-5 | F | Fever, Cough, Rhinorrhea, Wheezing | Bronchiolitis |
| 3-5 | M | Fever, Cough, Rhinorrhea, Sore Throat | Bronchiolitis |
| 3-5 | M | Fever, Cough, Rhinorrhea, Wheezing | Pneumonia |
| 3-5 | M | Fever, Cough, Rhinorrhea, Wheezing | Pneumonia |
| 6-15 | F | Fever, Cough, Sore Throat, Headache | URTI |

Note: M: Male, F; Female, URTI: Upper respiratory tract infection

### Seasonal variation of HMPV

35 samples were collected in January. Similarly, 52 and 18 samples were collected during the second (February) and third month (March) respectively. The incidence of HMPV was high in late winter followed by early spring. The positive rate of HMPV was 14.3% during January followed by 13.5% and 11.1% in February and March month respectively (Figure 1).

**Figure 1:**
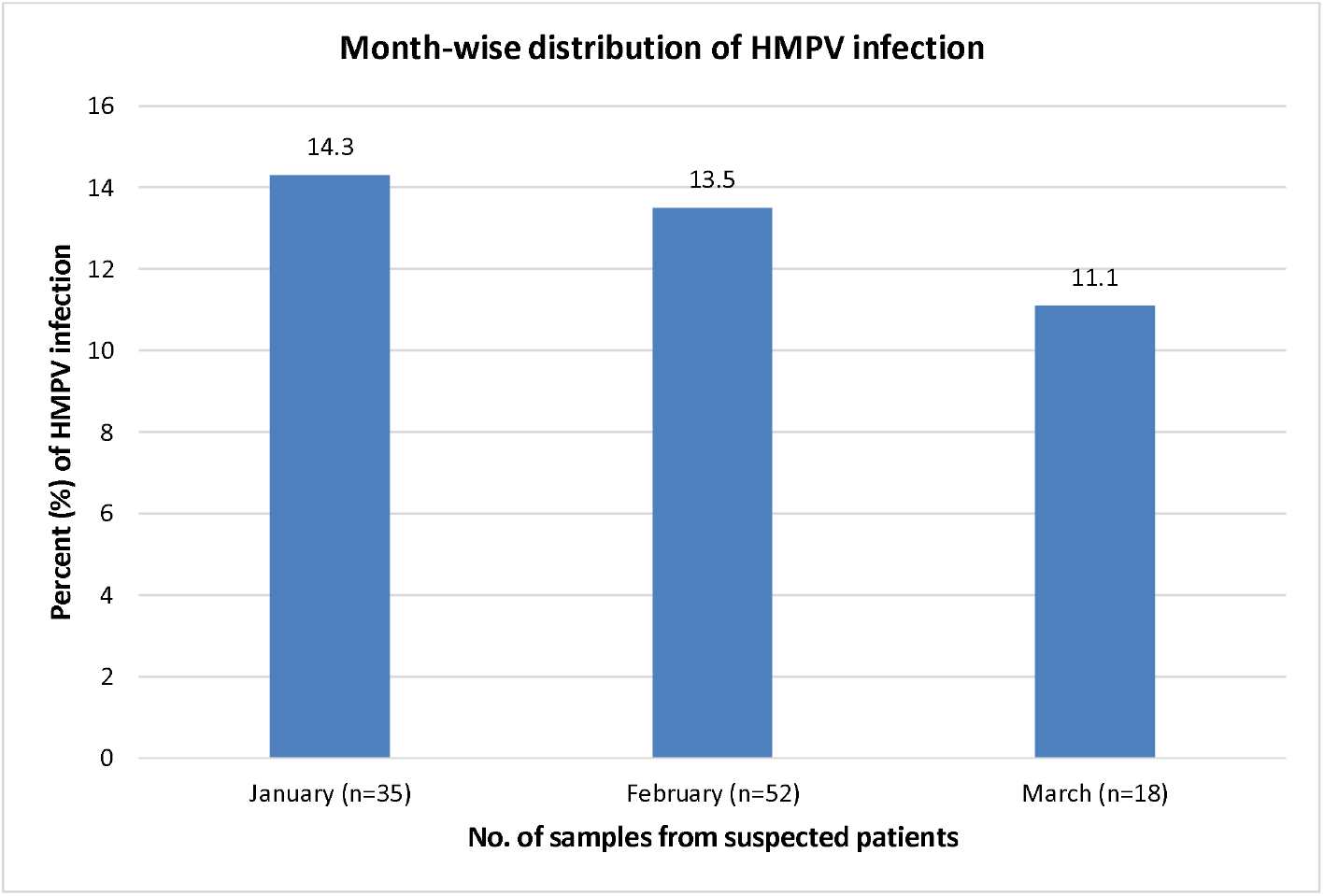
Month-wise distribution of HMPV infection among suspected children.

### HMPV co-infection with other viruses

Among 14 HMPV positive samples, co-infection was seen in two different patients. The coinfection with Parainfluenza virus was detected in 7.1% (1) of patients age group <3 and in 7.1% (1) of the patient with RSV in of age group 3-5 years. No co-infection was observed in patients in the age group 6-15 years. Both co-infection cases were found in children in the outpatient department (Table 5).

**Table 5:** Co-infection of HMPV with other bacteria and viruses among different age groups.

| Age group | Total positive sample | Co-infection |  | Total |
| --- | --- | --- | --- | --- |
|  |  | RSV (%) | PIV (%) |  |
| <3 | 7 | 0 | 1 (7.1%) | 1 |
| 3-5 | 6 | 1 (7.1%) | 0 | 1 |
| 6-15 | 1 | 0 | 0 | 0 |
| **Total** | **14** | **1** | **1** | **2** |

## DISCUSSION

Since 2001, after the discovery of HMPV, several studies have provided important information on HMPV epidemiology, clinical symptoms associated with HMPV infection, and the patient groups that are at risk for HMPV infection (van den Hoogen, 2003; van den Hoogen et al 2001). Although few studies have suggested that HMPV is an important causative pathogen in children of Nepal (Mathisen et al 2010), the actual data regarding HMPV infection in the country’s population remains largely unknown due to the limitations in these studies. To our knowledge, the result of this study is the first to report the incidence of HMPV in Nepal in children below the age of 15. Prior to this investigation, only studies regarding RTIs examined the role of HMPV along with other causative agents. In this study, we describe the incidence of HMPV, co-infections with other viruses, and clinical presentation of HMPV, in a Kanti Children’s hospital setting, and compared the clinical characteristics among those HMPV positive cases.

HMPV was found in 13.3% of children with ARIs. Different studies from different parts of the world have reported variable frequencies of HMPV infection, ranging from 2.2% to 43% (Maggi et al 2003; Aberle et al 2008; Lu et al 2011), with young children being the main groups in which HMPV infections are detected. Comparable to other reports (Banerjee et al 2007; Miroballi et al 2010), this study showed the highest incidence of HMPV in children below the age of three years among the study population. The study also confirms that HMPV is one of the major causes of ARIs in children of Nepal.

The prevalence of HMPV in children of age group <3 years was found to be 21.8%, followed by the prevalence of age group 3-5 years and 6-15 years at 13.3% and 3.5% respectively. The high prevalence of HMPV infection in little children might be attributed to weak immune function. According to NREVSS in a year-round survey that included all age groups from 2004 to 2008, 3.6% of specimens were positive for HMPV among consistently reporting laboratories in the United States. However, in this study, the positive rate of HMPV was 13.3%. The high prevalence might be because the study was done during the winter and spring season and only includes children (Busses et al 2010).

No HMPV-positive children aged 6 to 15 years were hospitalized. Rates of HMPV-associated hospitalization are highest among children <3 years old. A similar result was reported by (Honda et al 2006). The HMPV infection in younger children is more severe as compared to adults probably due to the under-developing immune system.

In this study, we found the most common clinical symptoms of HMPV-associated with ARTI were fever, cough, rhinorrhea, sore throat, and wheezing. Among the 14 HMPV-positive children, cough, fever, rhinorrhea, and sore throat were the most common features Wheezing was present in approximately half of the children. No Earache and posttussive emesis were found. Comparable data was reported in Honda et al (2006) study. Previous studies indicated that HMPV causes upper and lower RTIs in patients of all ages, but mostly in children aged below 5 years (Falsey et al 2003; van den Hoogen et al 2003).

Clinical diagnoses of the HMPV-positive children in this study included upper respiratory tract infection (14.3%), pneumonia (42.9%), bronchiolitis (28.5%), and asthma (14.3%). Several reports indicate that HMPV is a commonly identified cause of pediatric lower RTIs, and is second only to RSV as a cause of bronchiolitis in early childhood (van den Hoogen et al 2003). A large epidemiological retrospective study carried out over 20 years, detected HMPV in 1%-5% of pediatric upper RTIs (UTRIs), with variation from year to year (William et al 2006). They reported bronchiolitis as the most common presentation of HMPV illness. However, in this study, pneumonia was the most common clinical diagnosis and there were no cases of otitis media. For more accurate data sample size and study period must be increased.

The seasonal distribution of HMPV infection varies. Some longitudinal studies (Banerjee et al 2007; Nidaira et al 2012) suggested that the high season for HMPV is from winter to spring (between December and May) and the low season is the fall (around September and October). This study was carried out in the late winter to the early spring season and the prevalence was 13.3%. As Kathmandu has a subtropical climate it is likely the highest season for HMPV. However, the high season for HMPV in tropical and subtropical areas varies from winter to spring in Brazil, spring and/or summer in Taiwan, and the rainy season in Vietnam (Nidaira et al 2012). During the three-month study period of this research, the highest rate of HMPV was in January followed by February, and dropped to a minimum during March. Comparable results were reported from Norway. HMPV appeared to be peaking in the winter months. Smaller outbreaks occurred during the spring and early summer months and coincided with a reduction in the total number of children admitted with RTIs (Ditt et al 2011). Although the reasons are not yet known, this trend may differ from that of other tropical and subtropical areas. However, our investigation period was short. Thus, to clarify the prevalence season of HMPV in Nepal, more longitudinal studies may be needed. Samples from different areas should be tested.

Due to the similar seasonal distribution of HMPV and other respiratory viruses, the potential co-infection likely existed. Some studies have found a co-infection rate of up to 70% in HMPV (Semple et al 2005; Calvo et al 2008). In this study, the majority of HMPV infected cases did not show any co-infections. Only two out of 14 positive patients were co-infected with other viruses. The co-infected viruses included respiratory syncytial virus and parainfluenza virus. Nevertheless, the lack of other respiratory pathogens in most patients suggests that HMPV is a true pathogen of both the upper and lower respiratory tracts.

As a major study limitation, we couldn’t conduct our study throughout the year including all seasons because of a limited budget and time. Therefore, the generalization of the findings may not be precise as the study included a low number of cases collected in three months. Further study for an extended period including more populations from different groups is recommended to generate the baseline information to address the magnitude of the disease and rational use of antibiotics therapy.

## CONCLUSION

From these studies, it can be concluded that HMPV is an important human pathogen associated with RTI in young children, immune-compromised individuals, elderly individuals, and, to a lesser extent, other populations. In addition, this study was the first to report HMPV as an important cause of acute RTI in children under 5 years of subtropical countries like Nepal using multiplex real-time RT-PCR. HMPV can cause a wide range of respiratory tract illnesses ranging from minor upper respiratory tract to severe bronchiolitis including pneumonia and asthma. The severity is higher in young children as compared to older children often leading to hospitalization. Likewise, HMPV co-infection with RSV and Para influenza virus was also reported in our study indicating HMPV infection as one of the predominant causes of ARI in children in Kathmandu. The study finally suggests immediate care in case of the flu-like syndrome among children to avoid the potential respiratory severity of HMPV.

## Data Availability

All data produced in the present study are available upon reasonable request to the corresponding author.

## Abbreviations

ARI: Acute Respiratory Tract Infection
CDC: Center for Disease Control and Prevention
cDNA: Complimentary DNA
COPD: Chronic Obstructive Pulmonary Disease
Ct: Cycle threshold value
DNA: Deoxyribonucleic acid
HMPV: Human Metapneumovirus
NC: Negative control
PC: Positive Control
RTIs: Respiratory Tract Infections
RNA: Ribonucleic acid
RSV: Respiratory Syncytial Virus
RT-PCR: Reverse Transcriptase Polymerase Chain Reaction
URTIs: Upper Respiratory Tract Infections
VTM: Viral Transportation Media.

## Author Contributions

All authors had substantially contributed to the conception and design of the study, acquisition of data, or analysis and interpretation of results and equally took part in drafting the manuscript or revising it. All authors had read and approved the final version of the manuscript and agreed to submit it to this journal.

## Institutional Review Board Statement

Ethical approval was obtained from the Institutional Review Committee of Kanti Children’s Hospital, Maharajgunj, Kathmandu, Nepal (Reference no.: 2018019).

## Informed Consent Statement

Patient consent and assent was obtained before the collection of specimen and data from respective parents or local guardians of patients.

## Data Availability Statement

The raw data of the study will be available on request to the corresponding author at.

## Acknowledgments

We would like to thank all the supporting staff of the Department of Microbiology, GoldenGate International College, Battisputali; Kanti Children’s Hospital, Maharajgunj, and Central Diagnostic Laboratory & Research Center, Kamalpokhari, Kathmandu, Nepal

---

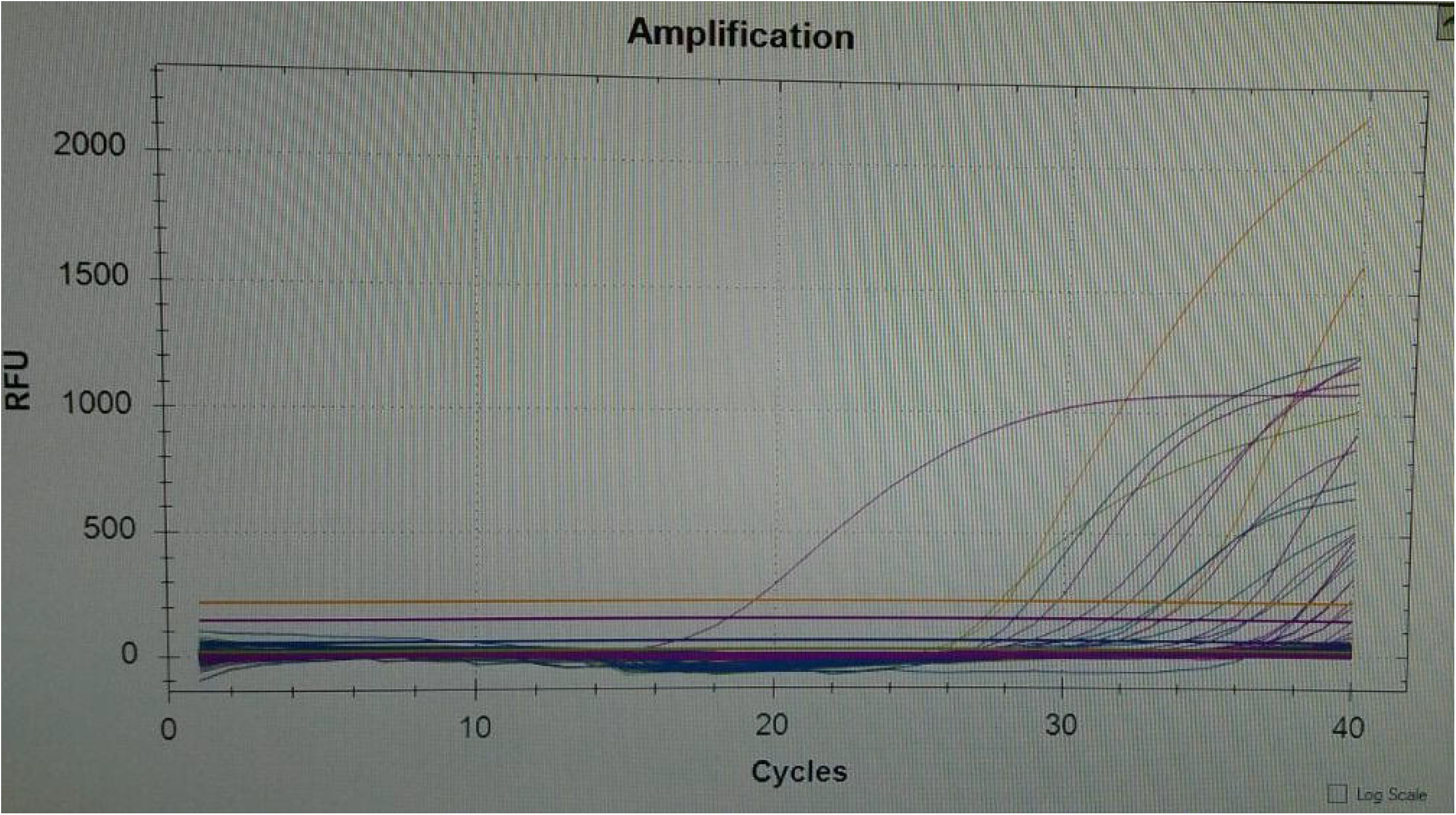

